# Characterization of Leukodystrophy Penetrance Using Large-Scale Integration of Genomic Population Screening and Electronic Health Records

**DOI:** 10.64898/2026.07.04.26356970

**Authors:** Hannah C. Happ, G. Bryce Christensen, Stacey Knight, Alfredo Novoa, Darin Isakson, Lincoln D. Nadauld, Aaron R. Quinlan, Joshua L. Bonkowsky

## Abstract

**Background and Objectives:** Leukodystrophies are rare genetic diseases affecting the central nervous system white matter, leading to progressive disabilities and death. Although early diagnosis is critical for therapies, the penetrance and phenotypic spectrum of many leukodystrophies remain poorly defined. Here, we integrate sequencing population screening with longitudinal electronic health record (EHR) data. Our goals were to assess the prevalence of undiagnosed leukodystrophy, characterize phenotypic variability among genotype-positive individuals, and estimate penetrance across multiple leukodystrophies.

**Methods:** We analyzed 19 genes associated with 13 leukodystrophies in pediatric and adult individuals recruited via the HerediGene Population Study, a 5-year study conducted primarily of healthy individuals in the U.S. intermountain west. Sequencing was performed on 210,983 individuals, consisting of genome sequencing for 34,033 and SNP panel imputation for 176,950. Variant results were cross-referenced to comprehensive and longitudinal (20+ years) clinical data in the Intermountain Health Enterprise Data Warehouse and to the Utah Leukodystrophy Program.

**Results:** Pathogenic variants were identified in 4 genes (*CSF1R, PLP1, POLR3A, SNORD118*) in 9 individuals, none of whom had a clinical leukodystrophy diagnosis or characteristic MRI findings. These findings suggest that missed clinical diagnoses of most leukodystrophies are uncommon in a centralized healthcare system, but also demonstrate that for some leukodystrophies there may be variable or reduced penetrance, or broader phenotypic spectra than recognized. We used published incidence estimates and the observed leukodystrophy-associated genotypes to infer penetrance ranges that varied from wide for ultra-rare leukodystrophies, to tightly bounded for more prevalent conditions.

**Discussion:** In this predominantly healthy population, we did not find any patients with leukodystrophy who had been genetically undiagnosed but then identified by sequencing. However, we identified 9 individuals with genotypes previously reported to result in leukodystrophy, but none of whom had clinical symptoms or MRI features associated with the specific leukodystrophy. Our results support a revised model in which leukodystrophies exist along a continuum of penetrance and expressivity, with implications for newborn screening, variant interpretation, and risk stratification.

## Introduction

Leukodystrophies are a group of genetic diseases affecting the white matter of the central nervous system in children and adults, leading to progressive disabilities and death over months to years (Bonkowsky et al., 2010; Gordon et al., 2014; Vanderver et al., 2015; Bonkowsky et al., 2021). While individually rare, collectively there is an incidence of greater than 1 in 4,500 live births (Bonkowsky et al., 2010; Soderholm et al., 2020). There are more than 400 different genes known to cause leukodystrophy (Lynch et al., 2017; Urbik et al., 2020); treatments are only available currently for a few, but multiple clinical trials are underway with promising advances (Ceravolo et al., 2023; Metovic et al., 2024; Nagy et al., 2026). Treatment is dependent on timely diagnosis: delayed or failure of diagnosis is associated with increased mortality, and for some leukodystrophies there is a narrow time window of treatment efficacy (van der Knaap et al., 2019; Page et al., 2019; Ghabash et al., 2021; Wolf et al., 2025).

A central unresolved question in leukodystrophy work is the true penetrance and expressivity of disease. Classical descriptions have emphasized high penetrance and severe phenotypes; however, multiple lines of recent evidence challenge this paradigm (Brimley et al., 2013; Saavedra-Matiz et al., 2016; Takano et al., 2019; Elgun et al., 2019; Trinidad et al., 2023; Lund et al., 2026). This also raises the possibility that some individuals with leukodystrophy might be mis- or underdiagnosed (Bonkowsky et al., 2018; Bonkowsky et al., 2024; Mohajer et al., 2025). There are several lines of evidence for these concerns. First, predicted pathogenic/likely pathogenic (P/LP) variants in leukodystrophy genes have been identified, including in newborn screening, gnomAD, and other sources, but which have not been associated with disease (Soderholm et al., 2020; Trinidad et al., 2023; Gagliardi et al., 2025). Second, recent expansion of newborn screening, such as for X-linked Adrenoleukodystrophy (ALD), has identified many individuals with predicted P/LP *ABCD1* gene variants *and* biochemical involvement consisting of elevated very long chain fatty acids (VLCFAs), but no presence of clinical symptoms, even in multi-generation pedigrees (Baker et al., 2022; Onuki et al., 2025; Lund et al., 2026). Third, even with the same genetic mutation, different individuals have different leukodystrophy presentations, suggesting the occurrence of genetic modifiers (Wiesinger et al., 2015; Wichers et al., 1999; Takano et al., 2019).

Despite these insights, systematic population-based evaluations of penetrance integrating genomic and longitudinal clinical data remain limited. Here, we leverage a uniquely integrated dataset combining population-scale genomic screening with longitudinal EHR data and a regional leukodystrophy registry to (1) assess the prevalence of undiagnosed leukodystrophy, (2) characterize phenotypic variability among genotype-positive individuals, and (3) estimate penetrance across multiple leukodystrophies. This was performed in the intermountain west region of the U.S., in which health care and follow-up are centralized allowing unique ascertainment capability. Findings from this project are critical not only for understanding leukodystrophy pathophysiology but also for informing newborn screening policies, variant classification frameworks, and therapy delivery.

## Methods

### Standard Protocol Approvals, Registrations, and Patient Consents

This study was approved by the Intermountain Healthcare and University of Utah Institutional Review Boards (IRBs); all participants provided written informed consent.

### Study Design and Data Sources

This was a population-based genetic study, with retrospective review of clinical and demographic data.

### Study Patients

Participants consisted of the HerediGene Population Study cohort (Taylor et al., 2024), which recruited individuals seen at an Intermountain Healthcare (IH) facility. IH is a healthcare system that consists of 33 hospitals and 385 clinics in Utah and neighboring states. IH is a regional, not-for-profit integrated health care delivery system, with 33 hospitals, and 385 clinics and urgent care facilities located across the region, serving 60% of Utah’s 3.4 million residents and 85% of the state’s children (Byington et al., 2012; James and Savitz, 2011; U.S. Census Data 2022). The children’s hospital (Primary Children’s Hospital) and affiliated IH and University of Utah pediatric specialty clinics serve as the sole tertiary pediatric care for an estimated pediatric population of >1.7 million children (Annual Estimates, 2022), and the major site of pediatric specialty care in the region. The pediatric neurology group at Primary Children’s Hospital/University of Utah, including the leukodystrophy specialist (Utah Leukodystrophy Program, see below), serve as the only source of pediatric neurology care in Utah and Wyoming and as the predominant source for the intermountain region.

Participants were approached in hospital or clinic settings; health fairs; letter and phone contacts; and through self-referral. Inclusion was any individual, including healthy or diagnosed with a medical condition, and they were not required to have insurance or to be a patient of Intermountain Healthcare. (See ***Supplemental Table 1*** for full inclusion/exclusion criteria.) Samples included collection from buccal cheek swabs, blood draws, and residual blood from routine lab tests. Written consents were obtained in person or via e-consenting; for pediatric participants a parental consent was obtained with assent as appropriate. The participant had to live in the United States and legal authorization representative consents were not allowed. Reporting of ACMG and selected gene variant secondary findings are being performed as part of this study.

### Sequencing Program

Whole-genome DNA sequencing and genotyping was performed in parallel at IH Intermountain Precision Genomics (IPG) laboratory in St. George, Utah and at deCODE Genetics in Iceland following protocols described previously (Gudbjartsson et al., 2015). Whole genome sequencing (WGS) was performed on 34,033 participants, using NovaSeq Illumina technology. All participants were genotyped at IPG or deCODE using the Illumina Global Screening Array platform. Whole genome imputation analysis was performed at deCODE for all participants using a multi-ethnic haplotype reference panel derived from approximately 60,000 genomes including all sequences from the current study as well as additional genomes from other populations primarily of European descent (Thorolfsdottir et al., 2024). Over 245 million high-quality sequence variants and indels, sequenced to a mean depth of 20×, were identified via joint genotyping using Graphtyper (v.2.7.5) (Eggertson et al., 2017), and phasing was carried out using SHAPEIT (O’Connell et al., 2014). Quality-controlled chip genotype data were phased using Shapeit 4 (Deleneau et al., 2019). After imputation and quality checking, a total of 210,983 participants had imputed and/or sequenced genetic results.

Sequences were analyzed for likely pathogenic variants in the leukodystrophy genes outlined in ***Supplementary Table 2*** and ***Supplementary Table 3***; and ClinVar numbers and specific variants are listed.

### Clinical Data

Demographic information, ICD-coded diagnoses, laboratory and radiology results, and clinical data, were retrieved from the IH Enterprise Data Warehouse (EDW). The EDW records all clinical, pharmacy, laboratory, and radiological outcomes for all IH patients, including more than 10 Terabytes and 9,000+ data tables (Evans et al., 2012; Nelson et al., 2013). Race/ethnicity status stored in the EDW is self-reported at the time of clinical presentation.

Manual review of individual cross-referenced patient clinical information was subsequently performed by a leukodystrophy specialist (JLB). Participants with predicted P/LP variants in leukodystrophy genes were also cross-referenced with the Utah Leukodystrophy Program (ULP) database (Bonkowsky et al., 2010), which consists of >20 years of data for all leukodystrophy patients (adult and pediatric) in the intermountain west region; there is no other leukodystrophy specialist or referral site in the region.

### Statistical Analysis

Descriptive statistics were used to characterize the study cohort with data presented as frequency and percent, unless otherwise specified. Deceased participants (15,737) and those over age 90 (3,177) were removed from the age analysis. Analyses were conducted in GraphPad Prism 9; an alpha of 0.05 was used to assess statistical significance.

Figure 1 graphics generated with assistance of BioRender.

**Figure 1.**
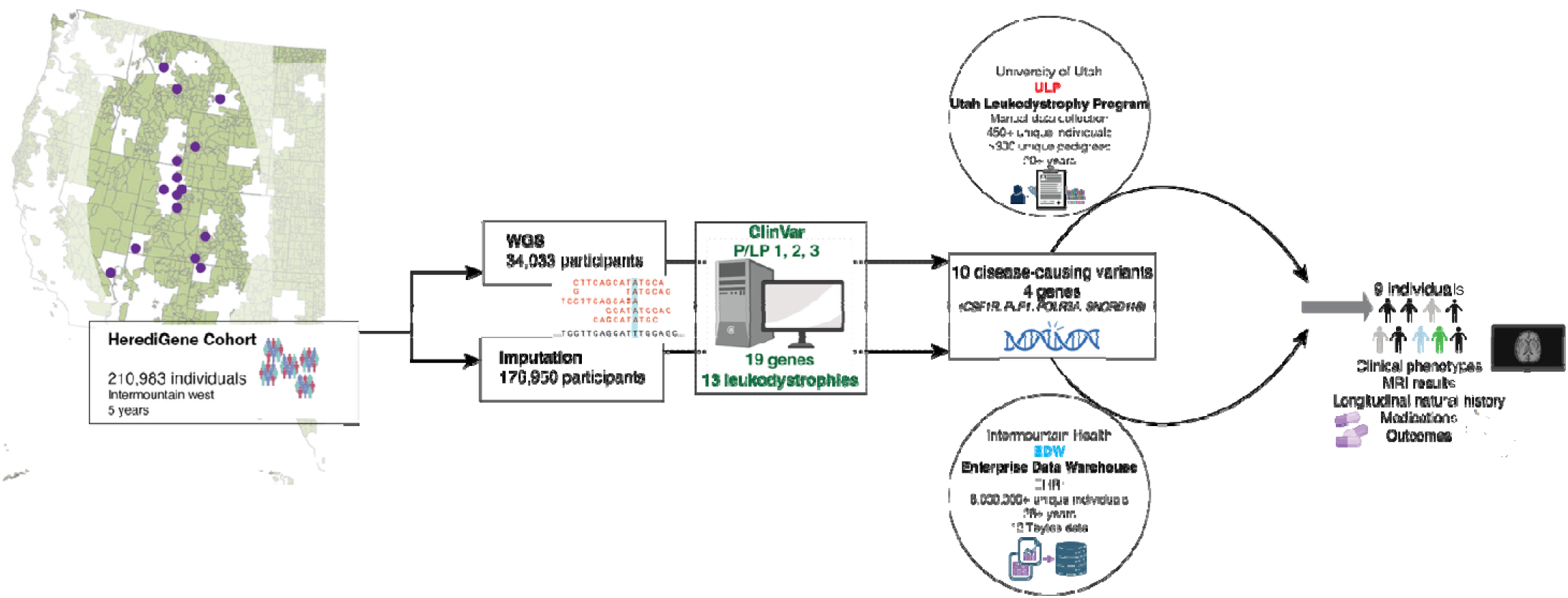
Overview of cohort development, analytical approach, and leukodystrophy genes.

### Determination of Penetrance

Penetrance was defined as the probability of disease given a pathogenic genotype, **π = P(D = l|G = l)**, where ***D*** is disease status and ***G*** is the presence of a P/LP genotype consistent with the associated LD. To account for uncertainty in the observed count, for each LD with a published incidence estimate ***I***, we modeled the frequency of disease-associated genotypes among unaffected individuals (defined according to MOI) as a binomial random variable to estimate a 95% confidence interval for that frequency ***q***. Penetrance was estimated using the equation 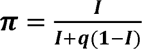. Where high and low incidence ranges were available, lower and upper bounds of penetrance were obtained using the lower incidence estimate and upper confidence limit of ***q*** or upper incidence estimate and lower confidence limit of q, respectively. When the lower bound of ***q*** was 0, the upper penetrance bound was reported as 1.

### Data availability

Anonymized data not published within this article will be made available by request from a qualified investigator and with approval from deCode/Amgen.

## RESULTS

Participant identification, genome sequencing and imputation, and cross-integrations with the electron c health record and with the regional leukodystrophy database are shown in ***Figure 1***. Participants consisted of adult and pediatric individuals living in the intermountain west region, who enrolled in the HerediGene Population Stud, a 5-year prospective screening study conducted primarily for non-diagnosis purposes of healthy individuals. The final cohort consisted of 210,983 individuals. Deceased participants (15,737) and those over age 90 (3,177) were removed from the age analysis. The cohort had a mean age of 54.4 years and had more females than males (60% versus 40%, respectively) (***Table 1***). 90% of the cohort was white, 7% did not report or declined to report race, and there were fewer than 3% of other racial groups represented. The cohort was 6% Hispanic, 88% non-Hispanic, and the remaining 6% did not report or declined to report on Hispanic ethnicity.

**Table 1.** Cohort demographics. WGS participants had both whole genome sequencing and imputation performed. Imputation participants had only imputation performed. Deceased participants (15,737) and those over age 90 (3,177) were removed from age analysis. Percentages of 1% or less not listed.

| 369 | 370 | Overall (210,658) | WGS (33,770) | Imputation (176,918) |
| --- | --- | --- | --- | --- |
| 371 | Characteristic |  |  |  |
| 372 |  |  |  |  |
| 373 | Age (years) |  |  |  |
| 374 | Mean | 54.4 | 61.0 | 53.3 |
| 375 | Median | 55.6 | 65.4 | 53.7 |
| 376 | Mode | 73.1 | 73.0 | 71.2 |
| 377 | Range | 2.5 – 90 | 2.7 – 90 | 2.5 - 90 |
| 378 |  |  |  |  |
| 379 | Sex |  |  |  |
| 380 | N (%) |  |  |  |
| 381 | Male | 83,246 (40%) | 15,108 (45%) | 68,154 (39%) |
| 382 | Female | 127,400 (60%) | 18,660 (55%) | 108,754 (41%) |
| 383 | Not reported or |  |  |  |
| 384 | Reported non-binary | 13 | 2 | 12 |
| 385 | Race |  |  |  |
| 386 | N (%) |  |  |  |
| 387 | White | 188,988 (90%) | 30,050 (89%) | 158,963 (90%) |
| 388 | Unavailable | 9,495 (5%) | 1,823 (6%) | 7,674 (4%) |
| 389 | Patient declined | 4,556 (2%) | 612 (2%) | 3,947 (2%) |
| 390 | Asian | 2,264 | 261 | 2,003 |
| 391 | American Indian/<br>Alaska Native | 1,433 | 307 | 1,126 |
| 392 | Black | 1,340 | 230 | 1,110 |
| 393 | Hawaiian/Pacific<br>Islander | 1,308 | 315 | 993 |
| 394 | Multiple | 1,291 | 176 | 1,115 |
| 395 |  |  |  |  |
| 396 |  |  |  |  |
| 397 | Ethnicity |  |  |  |
| 398 | N (%) |  |  |  |
| 399 | Not Hispanic | 184,940 (88%) | 28,794 (85%) | 156,173 (88%) |
| 400 | Hispanic | 12,992 (6%) | 1,540 (5%) | 11,453 (7%) |
| 401 | Unavailable | 8,213 (4%) | 2,711 (8%) | 5,502 (3%) |
| 402 | Patient declined | 4,533 (2%) | 729 (2%) | 3,806 (2%) |
| 403 |  |  |  |  |

We analyzed 19 genes for 13 different leukodystrophies for pathogenic or likely pathogenic genotypes, including for diseases with X-linked, dominant, and autosomal recessive modes of inheritance (***Table 2***). Whole genome sequencing was performed on 34,033 participants, which was used to inform SNP panel-based imputation of an additional 176,950 individuals. We recorded variants that were classified as pathogenic (P) or likely pathogenic (LP) according to ClinVar (VCF download version 20250115). Pathogenic variant carriers were identified for most leukodystrophy genes, except *ABCD1*, *GFAP*, and *TUBB4A*. A complete listing of variants identified in the leukodystrophy genes of participants, including the number of different P/LP variants reported in ClinVar for each gene each level (tier) 1, 2, or 3, is provided in ***Supplemental Table 2*** and ***Supplementary Table 3*.**

**Table 2.** Identified leukodystrophy gene ClinVar pathogenic or likely pathogenic (P/LP) variants, levels 1-3. Mode, Mode of inheritance; R, recessive; D, dominant; X, X-linked. Variant(s) combinations with presumed pathological disease outcomes highlighted, and presented further in. Tables 3 and 4.

|  |  | WGS Results (n=34,033) |  |  | Imputation Results (n=210,983) |  |  |  |
| --- | --- | --- | --- | --- | --- | --- | --- | --- |
| Genes | Mode | Carriers | Homozyg | Compound Het | Imputed Carriers | Homozyg | Compound Het (trans) | Compound Het (cis) |
| AARS2 | R | 73 | 0 | 0 | 0 | 0 | 0 | 0 |
| ABCD1 | X | 0 | 0 | 0 | 0 | 0 | 0 | 0 |
| ARSA | R | 226 | 0 | 0 | 937 | 0 | 0 | 0 |
| CSF1R | D | 1* | 0 | 0 | 2* | 0 | 0 | 0 |
| DARS2 | R | 67 | 0 | 0 | 156 | 0 | 0 | 0 |
| EARS2 | R | 67 | 0 | 0 | 285 | 0 | 0 | 0 |
| EIF2B1 | R | 3 | 0 | 0 | 15 | 0 | 0 | 0 |
| EIF2B2 | R | 64 | 0 | 0 | 321 | 0 | 0 | 0 |
| EIF2B3 | R | 33 | 0 | 0 | 110 | 0 | 0 | 0 |
| EIF2B4 | R | 10 | 0 | 0 | 47 | 0 | 0 | 0 |
| EIF2B5 | R | 77 | 0 | 0 | 410 | 0 | 0 | 0 |
| GALC | R | 64 | 0 | 0 | 164 | 0 | 0 | 0 |
| GFAP | D | 0 | 0 | 0 | 0 | 0 | 0 | 0 |
| PLP1 | X | 1 | 0 | 0 | 0 | 0 | 0 | 0 |
| POLR1C | R | 28 | 0 | 0 | 482 | 0 | 0 | 0 |
| POLR3A | R | 283 | 0 | 0 | 1267 | 1 | 0 | 0 |
| POLR3B | R | 86 | 0 | 0 | 430 | 0 | 0 | 0 |
| SNORD118 | R | 304 | 0 | 1** | 1531 | 4 | 1** | 0 |
| TUBB4A | D | 0 | 0 | 0 | 0 | 0 | 0 | 0 |

\* The patient with a WGS variant in CSF1R was also detected in the imputation and is counted in both columns.
\*\*The patient with the compound heterozygous finding in SNORD118 was found in both WGS and imputation and is counted in both columns.

Pathogenic or likely pathogenic genotypes that would result in clinical disease were identified in 4 genes, *CSF1R* (CRL, CSF1R-leukodystrophy)*, PLP1* (PMD, Pelizaeus-Merzbacher Disease; SPG2, Spastic Paraplegia Type 2)*, POLR3A* (PRL, POLR3-Related Leukodystrophy), and *SNORD118* (LCC, Leukoencephalopathy with calcifications and cysts) across 9 individuals (***Table 3***). For the variants, 3 were identified from WGS, and 6 from imputation. All the identified variants or variant combinations have been previously published to result in clinical disease.

**Table 3.**
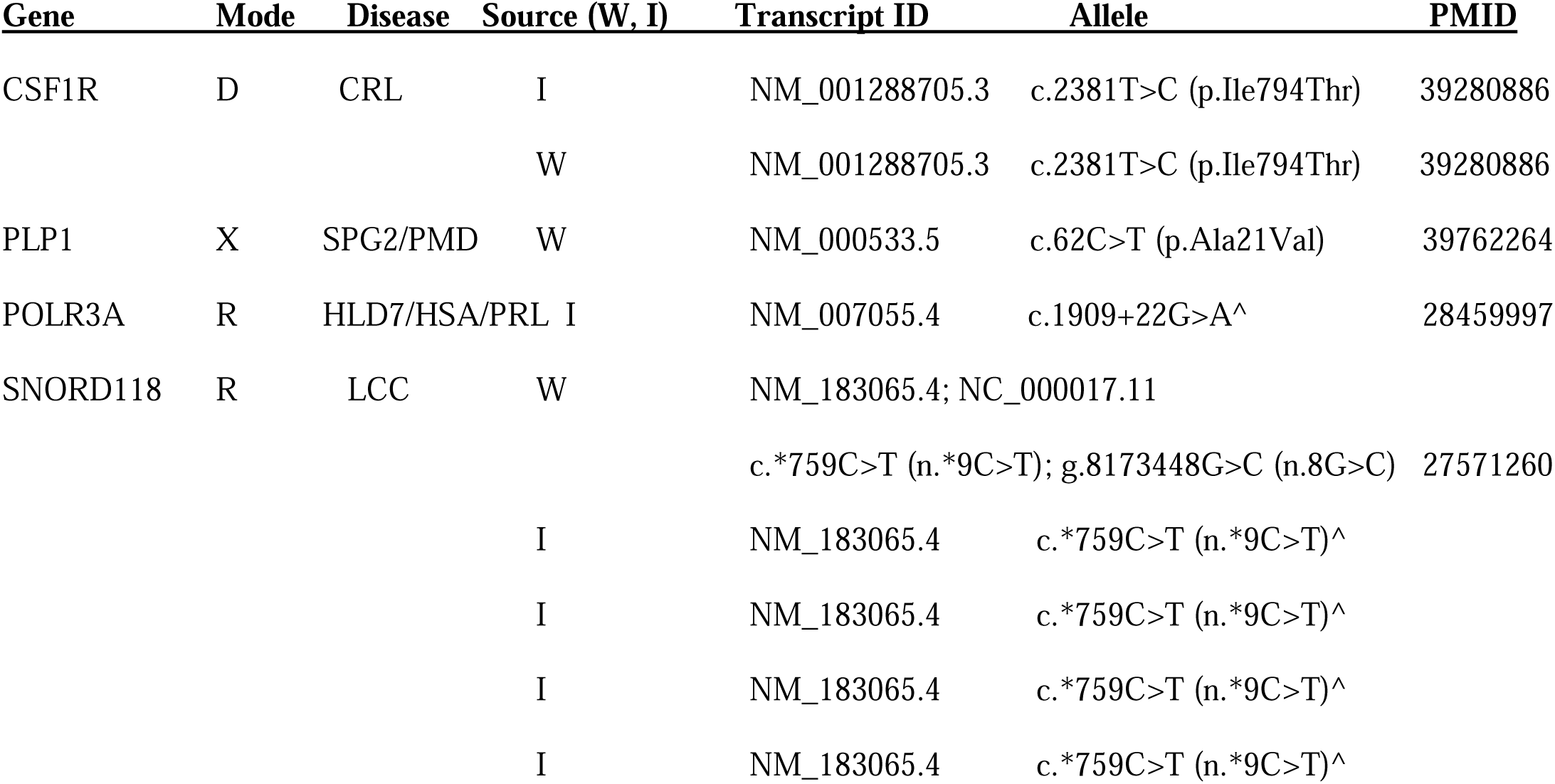
Variant details, pathogenic results. PMID refers to PubMed publication ID that reports leukodystrophy associated with the variant(s). Source, W= WGS; I = Imputation. CRL, CSF1R-leukodystrophy. HLD7, hypomyelinating leukodystrophy type 7. HSA, hereditary spastic ataxia. LCC, Leukoencephalopathy with calcifications and cysts. PMD, Pelizaeus-Merzbacher Disease. PMID, PubMed ID number (if any) reported with the variant; PRL, POLR3-Related Leukodystrophy; SPG2, Spastic Paraplegia Type 2. ^Homozygous for this variant.

| Gene | Mode | Disease | Source (W, I) | Transcript ID | Allele | PMID |
| --- | --- | --- | --- | --- | --- | --- |
| CSF1R | D | CRL | I | NM_001288705.3 | c.2381T>C (p.Ile794Thr) | 39280886 |
|  |  |  | W | NM_001288705.3 | c.2381T>C (p.Ile794Thr) | 39280886 |
| PLP1 | X | SPG2/PMD | W | NM_000533.5 | c.62C>T (p.Ala21Val) | 39762264 |
| POLR3A | R | HLD7/HSA/PRL | I | NM_007055.4 | c.1909+22G>A^ | 28459997 |
| SNORD118 | R | LCC | W | NM_183065.4; NC_000017.11 |  |  |
|  |  |  |  | c.*759C>T (n.*9C>T); g.8173448G>C (n.8G>C) |  | 27571260 |
|  |  |  | I | NM_183065.4 | c.*759C>T (n.*9C>T)^ |  |
|  |  |  | I | NM_183065.4 | c.*759C>T (n.*9C>T)^ |  |
|  |  |  | I | NM_183065.4 | c.*759C>T (n.*9C>T)^ |  |
|  |  |  | I | NM_183065.4 | c.*759C>T (n.*9C>T)^ |  |

For the 9 individuals with P/LP variants, we cross-referenced to unique patient clinical data (***Table 4***) to identify any clinical evidence of leukodystrophy. First, we analyzed whether any of the individuals had been evaluated or followed in the Utah Leukodystrophy Program (ULP). The ULP follows essentially all pediatric and adult leukodystrophy patients in the intermountain west region, with follow-up spanning retrospectively more than 20 years (Bonkowsky et al., 2010). None of the individuals were known or had been seen in the ULP.

**Table 4.** Clinical details for pathogenic variant results. ^, homozygous. MRI, brain MRI. TIA, transient ischemic attack. MRI; results: + indicates brain MRI performed; -, no MRI performed; “scattered T2” indicates scattered T2 signal hyperintensities. *Age range provided.

| Gene | Age* | Sex | Variant(s) | Symptoms/Diagnoses | MRI; results |
| --- | --- | --- | --- | --- | --- |
| CSF1R | 61-70 | F | c.2381T>C (p.Ile794Thr) | depression, migraines | +; scattered T2 |
|  | 31-40 | M | c.2381T>C (p.Ile794Thr) | depression | - |
| PLP1 | 91-100 | F | c.62C>T (p.Ala21Val) | - | - |
| POLR3A | 21-30 | F | c.1909+22G>A | - | - |
| SNORD118 | 81-90 | M | n.*9C>T; n.8G>C | kidney disease | - |
|  | 61-70 | M | n.*9C>T^ | polyneuropathy | - |
|  | 61-70 | F | n.*9C>T^ | - | +; scattered T2 |
|  | 71-80 | F | n.*9C>T^ | TIA; kidney disease | +; scattered T2 |
|  | 71-80 | M | n.*9C>T^ | kidney disease | - |

Second, we reviewed all clinical data for the individuals in the Intermountain Health Enterprise Data Warehouse (EDW), a comprehensive electronic medical record database with more than 28 years of longitudinal data. The EDW includes laboratory values, radiology reports, and clinic and in-patient notes, and both automated and manual chart review was performed. None of the 9 individuals with P/LP genotypes identified in this study were found to have a clinical diagnosis or symptoms typically associated with leukodystrophy. Further, none had brain MRI features of a leukodystrophy (***Table 4***). Ages ranged from 28 to over 90 years old. Only three individuals had brain MRIs performed obtained for other clinical indications, which showed minor scattered T2 signal hyperintensity findings. The clinical indications for MRI were because of headaches or migraines in two individuals, and because of a transient ischemic attack (TIA) in the other individual. Of the individuals with *SNORD118* pathogenic variants, which causes Leukodystrophy with Calcifications and Cysts (LCC), 3 of the 5 individuals had kidney disease, and 1 other had polyneuropathy. For the 2 individuals with *CSF1R* pathogenic variants, both had depression and were on anti-depressant medication treatment. The two individuals with *PLP1* or *POLR3A* variants had no clinical diagnoses and had only been seen for well visits.

Finally, we estimated the frequency *q* of pathogenic or likely pathogenic genotypes among unaffected individuals, and used these estimates, together with published estimates of leukodystrophy incidence, to infer plausible penetrance ranges across leukodystrophy conditions (***Table 5***). For several conditions with low reported incidence and no qualifying genotypes observed among unaffected individuals, the upper confidence limits of *q* is driven primarily by cohort size. In contrast, conditions with one or more P/LP genotypes among unaffected individuals yield more precise estimates of genotype frequency.

**Table 5.** Penetrance estimates of leukodystrophies. Incidence estimates based on published rates were compared to observed numbers in the cohort (N). A Credible Interval calculation was used to develop an Expected Counts range, and then used to calculate a range of disease Penetrance. Incidence refers to number per live births.

|  | Gene | Mode | Disease | N | Incidence Estimate |  | Observed | Credible Interval |  | Expected Counts |  | Penetrance |  |
| --- | --- | --- | --- | --- | --- | --- | --- | --- | --- | --- | --- | --- | --- |
|  |  |  |  |  | Low | High |  | Low | High | Low | High | Low | High |
| 466 | ABCD1 | X | ALD | 83,246 | 17,000 | 14,000 | 0 | 0.00E+00 | 4.43E-05 | 0 | 3.69 | 0.570 | 1.0 |
| 467 | ARSA | R | MLD | 210,658 | 112,700 | 40,000 | 0 | 0.00E+00 | 1.75E-05 | 0 | 3.69 | 0.336 | 1.0 |
| 468 | CSF1R | D | CRL | 210,658 | N/A | 3,500 | 2 | 1.15E-06 | 3.43E-05 | 0.24 | 7.23 | 0.893 | 0.996 |
| 469 | EARS2 | R | LTBL | 210,658 | N/A | 812,000 | 0 | 0.00E+00 | 1.75E-05 | 0 | 3.69 | 0.066 | 1.0 |
| 470 | EIF2B1-5 | R | VWM | 210,658 | 714,594 | 80,000 | 0 | 0.00E+00 | 1.75E-05 | 0 | 3.69 | 0.074 | 1.0 |
| 471 | GALC | R | Krabbe | 210,658 | N/A | 12,080 | 0 | 0.00E+00 | 1.75E-05 | 0 | 3.69 | 0.826 | 1.0 |
| 472 | GFAP | D | AxD | 210,658 | N/A | 2,700,000 | 0 | 0.00E+00 | 1.75E-05 | 0 | 3.69 | 0.021 | 1.0 |
| 473 | PLP1 | X | PMD/SPG2 | 83,246 | 750,000 | 52,632 | 1 | 3.04E-07 | 6.69E-05 | 0.03 | 5.57 | 0.020 | 0.984 |
| 474 | POLR3A,B;1C | R | PRL | 210,658 | N/A | 26,180 | 1 | 1.20E-07 | 2.64E-05 | 0.03 | 5.56 | 0.592 | 0.997 |

Penetrance estimates varied substantially across conditions. For disorders with extremely low reported incidence and no observed P/LP genotypes among unaffected individuals, penetrance was weakly constrained and the inferred ranges were broad, in some cases spanning nearly the full interval from 0-1. However, penetrance estimates were more constrained for conditions with relatively high incidence and/or with P/LP observed genotypes, which impose upper bounds on penetrance and reduce uncertainty (***Table 5***; ***Figure 2***).

**Figure 2.**
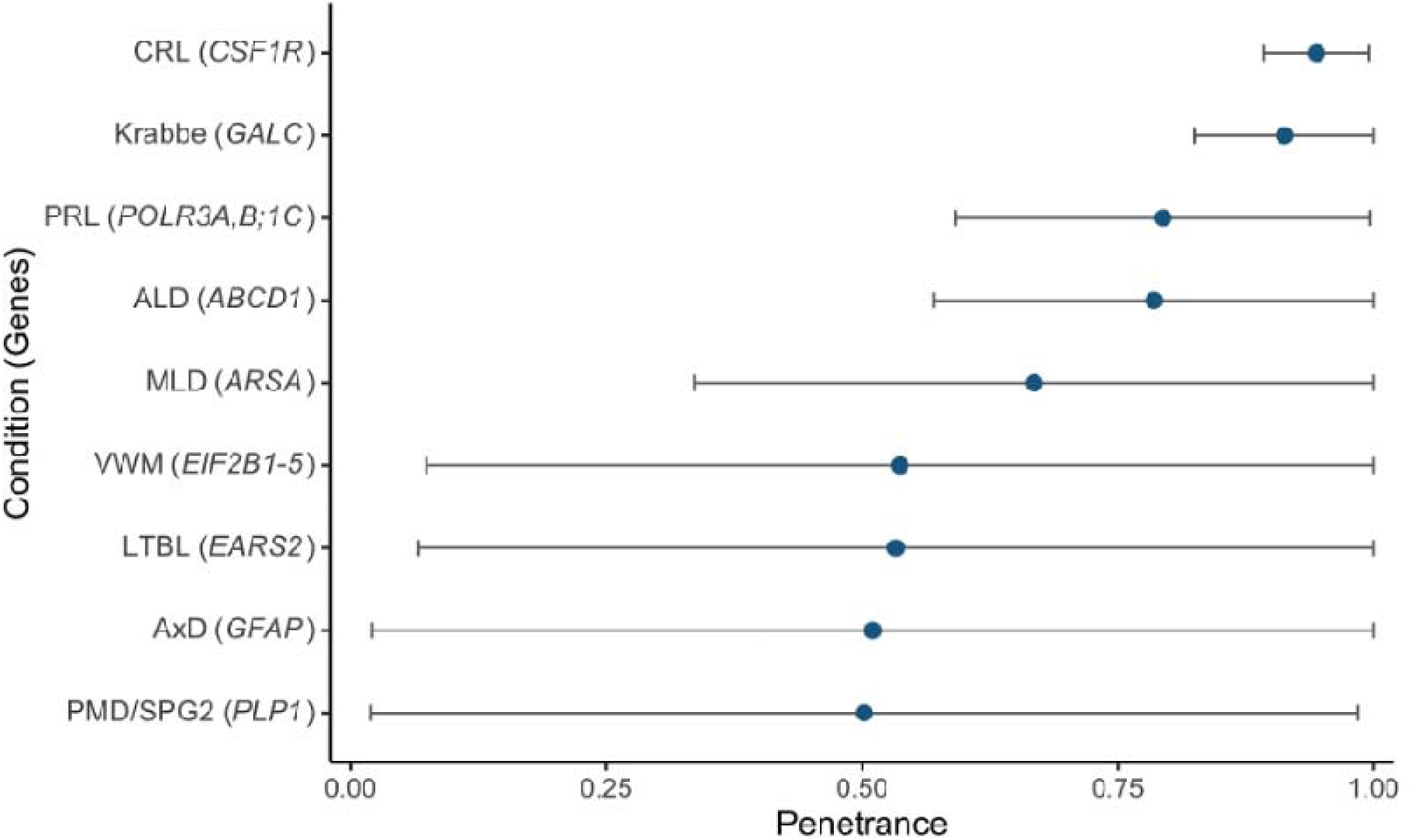
Leukodystrophy penetrance estimates, and 95% confidence intervals.

## DISCUSSION

In our work we have evaluated the extent of missed diagnoses, symptom variability, and disease penetrance, of leukodystrophies, which has not been systemically studied previously. This project is the largest of its type for leukodystrophies, and for which we combined population-based sequencing screening, using whole genome sequencing and SNP-based imputation, and integration with extensive longitudinal medical records as well as a unique comprehensive leukodystrophy database. In this predominantly healthy population, we did not find any patients with leukodystrophy who had been genetically undiagnosed but then identified by sequencing. However, we identified 9 individuals with genotypes previously reported to result in leukodystrophy, but none of whom had clinical symptoms or MRI features associated with the specific leukodystrophy.

Our findings support a shift away from a binary conception of penetrance toward a continuous, context-dependent model. The presence of individuals harboring previously reported pathogenic variants without clinical or radiographic evidence of disease suggests that penetrance may be modulated by additional factors beyond the primary genotype. Similar observations in population datasets, such as UK Biobank analyses of GFAP variants, indicate that reduced penetrance or subclinical disease may be more common than previously appreciated (Gagliardi et al., 2025). Importantly, this does not necessarily imply that variants are benign; rather, it suggests that disease expression may be delayed, attenuated, or organ-specific. Further, our findings show a broader variability in penetrance for leukodystrophy than previously recognized, and also revealed new insights into unexpected non-CNS phenotypes.

This study was carried out in the intermountain west region of the U.S., in which health care, particularly for specialty conditions such as pediatric neurology and leukodystrophy, are centralized to one or a few centers allowing unique ascertainment capability. This was particularly true for leukodystrophies, in which a single center (Utah Leukodystrophy Program) has provided more than 20 years of diagnosis and care for both children and adults, and is the referral center for newborn screening for the region, for a significant known majority of patients (Bonkowsky et al., 2010; Nelson et al., 2013). Further, with more than 200,000 individuals participating in the study, and because every individual was a patient of an integrated healthcare system with comprehensive and harmonized electronic medical records (Intermountain Health) used to track for clinical outcomes, we have high reliability for the clinical outcome conclusions.

Despite concerns as well as examples in the literature about missed or incorrect diagnosis (Bonkowsky et al., 2018; Mandia et al., 2025), we did not identify individuals with clinically manifest but genetically unrecognized leukodystrophy. This may reflect centralized specialty care; that the population for screening was primarily of healthy individuals; the longitudinal EHR depth; and the well-established referral pathways. In contrast, studies in less integrated systems demonstrate significant diagnostic delays (Mohajer et al., 2025), suggesting that health system structure is an important determinant for diagnosis.

For most of the leukodystrophy genes we studied (15) we did not find any individuals with clinically pathogenic variant combinations, consistent with the cohort development of the HerediGene study (Taylor et al., 2024). HerediGene was implemented as a healthy population screening project and not for diagnosis, although the study did not exclude individuals with medical diagnoses. For two of the genes with P/LP variants identified, *PLP1* and *POLR3A*, the individuals identified did not have any clinical symptoms. However, the *PLP1* individual was a female, and often females with *PLP1* variants have minimal or no symptoms (Wolf et al., 1999; Hurst et al., 2006).

Interestingly, both individuals with *CSF1R* pathogenic variants had depression, which has been reported in CRL (Rush et al., 2023; Wade et al., 2024). However, one of the two individuals had a brain MRI and was already 68 years old, suggesting an absence of CRL disease. The other individual was 34 years old, which can be a younger age for CRL than some people develop disease symptoms or findings on MRI.

For the individuals with *SNORD118* pathogenic variants, our findings suggest an expansion of the LCC disease phenotype to include kidney disease, as 3 of the 5 individuals had kidney disease. Although SNORD118 (LCC) has been found to cause kidney cysts in one patient (Bonomo et al., 2020), this publication was of a patient with extensive disease including of the brain. In contrast, our findings suggest a phenotypic expansion to possibly include isolated kidney disease.

Our data supports that a further evolution of understanding leukodystrophy symptoms, progression, and penetrance is necessary. First, it is increasingly clear that although many predicted pathogenic/likely pathogenic (P/LP) variants in leukodystrophy genes have been identified from a variety of sources, many have not been associated with clinical disease (Soderholm et al., 2020; Trinidad et al., 2023; Gagliardi et al., 2025). Further, even in patients in whom P/LP variants have been identified in individuals together with biochemical markers of disease, such as VLCFAs in ALD, galactocerebrosidase in Krabbe disease, or C16:0 sulphatides in MLD, but no disease symptoms, requiring further elucidation (Saavedra-Matiz et al., 2016; Trinidad et al., 2023; Lund et al., 2026). Thus, even in an individual with a known pathogenic variant accompanied by the biochemical marker of disease, and previously associated with clinical symptoms in other patients; prediction for that individual can show variability in timing of symptom onset, severity and progression of disease, and even in which symptoms he or she might have.

Our penetrance estimates had a broad range. For disorders such as PMD/SPG2 and AxD, penetrance intervals were necessarily broad because published incidence estimates are extremely low and penetrance cannot be meaningfully constrained; essentially we cannot distinguish between low penetrance and extremely rare incidence.

Here we would benefit from an even larger cohort. In contrast, several conditions have relatively high incidence estimates and penetrance ranges becomes more tightly constrained. We report the smallest penetrance range for CRL, a condition for which P/LP genotypes are observed among unaffected individuals and with a reported incidence of 1/3500. In this case, the estimated frequency of disease-associated genotypes is considerably lower than the incidence estimates, suggesting the majority of individuals with pathogenic genotypes develop disease. Under these conditions, penetrance is necessarily high; if penetrance were substantially lower, a much larger number of unaffected P/LP genotype carriers would be expected in the cohort. Thus, the combination of a relatively more common leukodystrophy (compared to other leukodystrophies in this study and the cohort size) and rare observation of pathogenic genotypes in a healthy cohort tightly constrains penetrance.

For conditions with a range of penetrance estimates, such as VWM and AxD, that include a plausible low or high penetrance, a true low penetrance could reflect a range of severity of disease caused by genetic or environmental modifiers or variable age of disease presentation and progression. Evidence for the former, that different variants have different penetrance or phenotype, is supported by variable natural history. For example, for VWM different variants and ages of onset are associated with different ages of disease presentation and speed of disease progression (Hamilton et al., 2018). However, there is also evidence for modifiers affecting phenotypes of disease. ALD is an example of this, in which even twin brothers with the same genotype have been reported to have discordant phenotypes (Korenke et al., 1996; Di Rocco et al., 2001).

Limitations in our study included that for determination of penetrance there are few established determinations of incidence (Bonkowsky et al., 2010; Vanderver et al., 2012; Soderholm et al., 2020; Hamilton et al., 2018), thus there was significant uncertainty in the incidence estimates. Further, some of the leukodystrophies in our cohort do not have any published incidence estimates, including LCC (*SNORD118*), TRL (*TUBB4A*), LBSL (*DARS2*), and ARD (*AARS2*). The majority of individuals in our cohort had a SNP panel and imputation performed, which may have resulted in false negative or false positive results. Imputation could not detect many structural variants or tissue-specific mosaicism, for example. The use of ClinVar for variant classification may result in absence of identifying pathogenic or likely pathogenic variants that have not been reported to ClinVar. We cannot exclude that some individuals might have later onset symptoms, or that individuals who did not have a brain MRI performed might in fact have CNS disease consistent with leukodystrophy if an MRI were performed, although their absence of clinical symptoms would argue against this possibility.

Leukodystrophies offer several important advantages for conducting this work and illustrate approaches that can be used for other genetic conditions. Particular strengths are that many leukodystrophies have distinct, specific, well-defined clinical phenotypes; are typically defined by the presence of abnormal brain MRI findings (van der Knaap and Valk, 2005; Vanderver et al., 2015; Urbik et al., 2020); and the presence of a multi-decade literature on natural history and on gene variants (Parikh et al., 2015; Kevelam et al., 2016; Mallack et al., 2022). Finally, there are now treatments and newborn screening for several leukodystrophies (Gordon-Lipkin and Fatemi, 2021; Ceravolo et al., 2024; Lin et al., 2025), providing an immediacy of relevance for our findings.

Additional work is needed to identify genetic and environmental modifiers of leukodystrophy penetrance. These modifiers will be important for understanding the pathophysiology of disease which in turn can drive more specific therapies. For example, if predictors of adrenal insufficiency could be identified in ALD (Kornbluh et al., 2024; Videbaek et al., 2025), this could in turn provide insight into how ALD is causing the adrenal insufficiency, and also for screening and treating that subgroup of ALD patients. Key next steps include integration of polygenic and modifier analyses, longitudinal follow-up of genotype-positive individuals, and functional validation of variants with uncertain penetrance. Insights can lead to individualizing treatment and management, moving away from a one-size-fits-all approach. Thus, while curative therapy would remain an ideal goal, more individualized treatment would reduce the over-medicalization risks of a leukodystrophy patient who might only develop a subset of symptoms.

Similarly, for newborn screening, understanding disease penetrance and risk is necessary to appropriately counsel families.

## ACKNOWLEDGEMENTS

JLB was supported by Primary Children’s Gene Kids; and NIH 1R01HD111570 and U54NS115052.

## Supplemental Data

**Supplemental Table 1.** Inclusion and exclusion criteria.

### Inclusion

- Male and female adults ≥ 18 years of age
- Children < 18 years of age (with or without a known suspected health and/or medical condition)
- Pregnant females (≥ 18 years of age)
- *<u>Any individual</u>* (with or without specific disease or injury, including Intermountain Health employees and study team members) who is a United States resident presenting to an Intermountain Health facility willing:
- to provide additional blood during a standard of care blood draw (for example, routine blood draw prior to surgery (inpatient or outpatient), during a routine clinic visit, during an inpatient encounter, during pregnancy or child birth, or presenting to an outpatient phlebotomy lab or clinic for a standard of care blood draw) <u>OR</u>
- to undergo a stand-alone blood draw procedure at an Intermountain Health facility <u>OR</u>
- provide residual blood (from an EDTA vacutainer) collected from a previously performed standard of care blood draw at Intermountain Health that will be otherwise discarded following the completion of ordered clinical laboratory tests.<u>OR</u>
- *<u>Any individual</u>* (with or without specific disease or injury, including Intermountain Health employees and study team members) who is a United States resident willing to undergo a stand-alone blood draw procedure at an Intermountain Health sponsored and/or affiliated event, outside of an Intermountain Health facility
- *<u>Any individual</u>* (≥ 18 years of age) presenting to the Gastrointestinal, Transplant/Liver, Pulmonary, or Orthopedic Specialty Group clinics at the Intermountain Medical Center (Intermountain Health, Salt Lake City, UT USA) or to The Orthopedic Specialty Hospital (Intermountain Health, Murray, UT USA) willing to provide an initial and one or more follow-up blood samples. Blood samples will be obtained during a routine clinic visit as either additional blood collected during a routine, standard of care blood draw procedure or as a stand-alone blood draw procedure at that clinic visit.
- *<u>Any individual</u> (*< 18 years of age) willing to provide a buccal swab at an Intermountain Health facility or at an Intermountain Health sponsored and/or affiliated event outside of an Intermountain Health facility.

### Exclusion Criteria

- Inability or refusal of the patient and/or the patient’s legally authorized representative to provide informed consent for any reason
- Inability or refusal of the child’s parent or legally authorized representative to provide parental permission for any reason

**Supplemental Table 2**

List from WGS and imputation sequencing results, reporting for each of the analyzed leukodystrophy genes the ClinVar variants identified in each tier; the number of carriers; and the number of homozygous or compound heterozygous individuals.

**Supplemental Table 3**

List of identified variants, sorted by pediatric and adult participants, for each of the analyzed leukodystrophy genes.

